# A Prospective, Randomized, Non-inferiority Clinical Protocol Comparing Single-port versus Multiport Extraperitoneal Robotic Radical Prostatectomy with the KangDuo Robotic System

**DOI:** 10.64898/2026.10.15.26365703

**Authors:** Tengfei Gu, Ting Chen, Yongtao Pan, Qinzhou Yu, Chen Wang, Jie Li

**Affiliations:** Department of Urology, Lishui Central Hospital (The Fifth Affiliated Hospital of Wenzhou Medical University), Lishui, China

**Keywords:** Prostate cancer, KangDuo robot, Single-port surgery, Multiport surgery, Extraperitoneal approach, Randomized controlled trial

## Abstract

**Background:** Robot-assisted radical prostatectomy is the standard minimally invasive treatment for localized prostate cancer. Conventional multiport robotic surgery is widely applied yet associated with more abdominal trauma and postoperative discomfort. Single-port robotic surgery can further minimize surgical injury. The domestic KangDuo robotic system has been gradually used in urological oncology, but high-level prospective evidence comparing single-port and multiport extraperitoneal radical prostatectomy remains scarce. This trial aims to compare the efficacy and safety of these two surgical approaches.

**Methods:** This is a single-center, prospective, 1:1 randomized, non-inferiority trial. Eligible patients diagnosed with cT1c–cT2cN0M0 localized prostate cancer will be randomized to receive either single-port (experimental group) or multiport (control group) extraperitoneal robotic radical prostatectomy using the KangDuo system. All participants will receive standardized enhanced recovery after surgery (ERAS) protocols. The primary endpoint is postoperative positive surgical margin rate. Perioperative parameters, postoperative recovery, medium-term functional recovery and oncological outcomes will be assessed comprehensively.

**Expected results:** We hypothesize that the single-port approach is non-inferior to the multiport approach in terms of oncological radical effect. The single-port technique is expected to provide better minimally invasive benefits, with comparable perioperative safety, urinary/sexual function recovery and mid-term oncological outcomes between groups.

**Conclusions:** This randomized trial will provide high-quality clinical evidence for the application of domestic single-port robotic surgery. The findings will facilitate individualized surgical selection for patients with localized prostate cancer.

## 1. Introduction

Prostate cancer is one of the most common genitourinary malignancies in elderly males, with a rising incidence in China. Robot-assisted radical prostatectomy has become the preferred minimally invasive treatment for clinically localized prostate cancer due to its superior three-dimensional vision, precise dissection and minimal tissue damage.

The extraperitoneal robotic approach avoids peritoneal cavity disturbance and accelerates postoperative intestinal recovery, which has been widely recognized in clinical practice. Nevertheless, traditional multiport robotic surgery requires multiple abdominal punctures, leading to increased wall trauma, postoperative pain and visible scars. With the development of ultra-minimally invasive surgery, single-port robotic technique emerges as a promising alternative, which preserves abdominal wall integrity and reduces surgical trauma via a single operative incision.

The KangDuo SR-2000 is a domestically developed surgical robot with favorable flexibility, stability and cost-effectiveness, and its application in urological tumor surgery is expanding. To date, there is a lack of prospective randomized controlled data comparing single-port and multiport extraperitoneal radical prostatectomy using the KangDuo system. The clinical value and suitable population of domestic single-port robotic surgery have not been fully clarified, which limits its standardized promotion.

Therefore, we designed this prospective randomized non-inferiority trial to compare intraoperative, postoperative and follow-up outcomes between the two surgical modalities. The results will offer evidence for the clinical application of domestic robotic systems and guide personalized treatment for patients with localized prostate cancer.

## 2. Study objectives and outcome measures

### 2.1 Primary objective and primary outcome

**①Primary objective**: To verify the oncological non-inferiority of single-port extraperitoneal KangDuo robotic radical prostatectomy compared with the multiport approach.

**②Primary outcome**: Postoperative positive surgical margin rate.

### 2.2 Secondary outcomes

**①Perioperative safety outcomes**: Total operative time, robotic docking time, estimated intraoperative blood loss, intraoperative transfusion rate, incidence of intraoperative visceral injury, 90-day overall and severe (Clavien-Dindo grade ≥ III) postoperative complication rates.

**②Postoperative recovery outcomes**: Pelvic drainage indwelling time, postoperative hospital stay, visual analog scale (VAS) pain score at discharge.

**③Medium-term functional outcomes**: Urinary continence rate (≤1 safety pad per 24 hours) and sexual function score (IIEF-5) at 3 and 6 months after surgery.

**④Oncological outcomes**: Prostate-specific antigen (PSA) control rate at 6 weeks postoperatively and 1-year biochemical recurrence-free rate.

## 2.3 Exploratory outcomes

Prespecified subgroup analyses will be performed according to tumor stage (T2a and below vs T2b/c), baseline PSA level (<20 ng/mL vs ≥20 ng/mL) and neoadjuvant therapy status.

Baseline functional scores and prostate volume will be analyzed as covariates to explore influencing factors of urinary and sexual function recovery.

## 3. Study design

This is a single-center, prospective, parallel-group, randomized controlled non-inferiority trial with a 1:1 allocation ratio. The whole study includes four phases: participant screening and enrollment, surgical intervention, postoperative management and long-term follow-up, and statistical analysis. All diagnostic, surgical and follow-up procedures are standardized to reduce confounding bias. The trial strictly follows the SPIRIT 2013 and CONSORT 2010 reporting guidelines.

## 4. Study setting and participants

### 4.1 Study setting

This trial will be conducted in the Department of Urology, Lishui Central Hospital (The Fifth Affiliated Hospital of Wenzhou Medical University). The surgical team has accumulated rich experience in KangDuo robotic operations and established standardized follow-up systems.

### 4.2 Inclusion criteria

① Male patients aged 18–85 years;

② Pathologically confirmed prostate adenocarcinoma via systematic prostate biopsy;

③ Clinical stage cT1c–cT2cN0M0 (8th AJCC TNM staging), without lymph node or distant metastasis;

④ Eligible for extraperitoneal robotic radical prostatectomy assessed by surgeons;

⑤ Eastern Cooperative Oncology Group (ECOG) performance status 0–1, tolerant to general anesthesia and robotic surgery;

⑥ Voluntary participation and signed written informed consent, with good compliance for long-term follow-up.

### 4.3 Exclusion criteria

① History of prostate ablation or radiotherapy (neoadjuvant endocrine therapy is allowed and recorded);

② Previous major pelvic or transperitoneal prostate surgery, or pelvic radiotherapy, which may disrupt extraperitoneal anatomy;

③ Imaging evidence of lymph node or distant metastasis;

④ Severe cardiac, pulmonary, hepatic, renal or coagulation disorders with anesthesia/surgery contraindications;

⑤ Uncontrolled acute or chronic infection;

⑥ Active malignant tumor within the past 5 years (stably controlled tumors excluded);

⑦ Mental disorders, cognitive impairment or substance abuse leading to poor compliance;

⑧ Other conditions judged inappropriate for enrollment by researchers.

### 4.4 Participant withdrawal and trial termination

#### Participant withdrawal

Voluntary withdrawal by patients or families; ineligibility reconfirmed after enrollment; intraoperative conversion of surgical approach; occurrence of severe adverse events; long-term loss to follow-up.

#### Trial termination

The trial will be prematurely terminated upon approval from the principal investigator and ethics committee in case of severe safety hazards, major protocol deviation or severely delayed recruitment.

## 5. Recruitment, screening and enrollment

Potential participants will be screened step by step according to inclusion and exclusion criteria. Qualified candidates will complete baseline assessment and sign informed consent before randomization. All recruitment procedures are standardized and traceable.

Researchers will fully inform participants of trial purpose, surgical schemes, potential risks and benefits. Informed consent will be signed voluntarily. All personal clinical data are strictly confidential and only used for this study.

## 6. Randomization and blinding

A web-based central block random system is used for 1:1 group allocation. Random sequences are generated by an independent biostatistician with strict allocation concealment. Stratified randomization is applied based on tumor stage and neoadjuvant therapy status to balance baseline characteristics.

Given the obvious differences in surgical appearance and operation, blinding of surgeons and patients is not feasible. Only the independent statistical analyst remains blinded to group allocation to avoid assessment bias.

## 7. Interventions

All patients receive unified ERAS perioperative management. Except for surgical approaches, all perioperative interventions are identical between two groups to eliminate confounding factors.

Unified perioperative protocol: No preoperative mechanical bowel preparation; oral carbohydrate intake 2 h before surgery; intraoperative goal-directed fluid therapy and normothermia management; postoperative multimodal analgesia and early ambulation; pelvic drainage removal when drainage volume < 50 mL; urinary catheter removal at postoperative 2 weeks; consistent anti-infection, fluid infusion and rehabilitation care. All operations are performed by the same senior surgical team.

### 7.1 Experimental group: Single-port extraperitoneal robotic radical prostatectomy

The KangDuo single-port robotic system is adopted. A 3–4 cm infraumbilical longitudinal incision is made to establish the extraperitoneal space. A single-port multi-channel device is placed, followed by pneumoperitoneum establishment and robotic docking. Standard procedures including bladder neck division, seminal vesicle dissection, Denonvilliers’ fascia incision, prostate resection, vesicourethral anastomosis are performed. A pelvic drainage tube is placed beside the single-port incision, and the incision is closed layer by layer.

### 7.2 Control group: Multiport extraperitoneal robotic radical prostatectomy

Conventional 3-arm plus 2-assistant port extraperitoneal robotic surgery is performed using the same KangDuo system. Patient position, anesthesia, operative approach, resection range, lymph node dissection indication, neuroprotection and anastomosis standards are fully consistent with the experimental group.

## 8. Data collection and follow-up schedule

All baseline, intraoperative, postoperative and follow-up data are recorded in standardized forms. Original medical documents are archived for traceability.

**Table 1.**
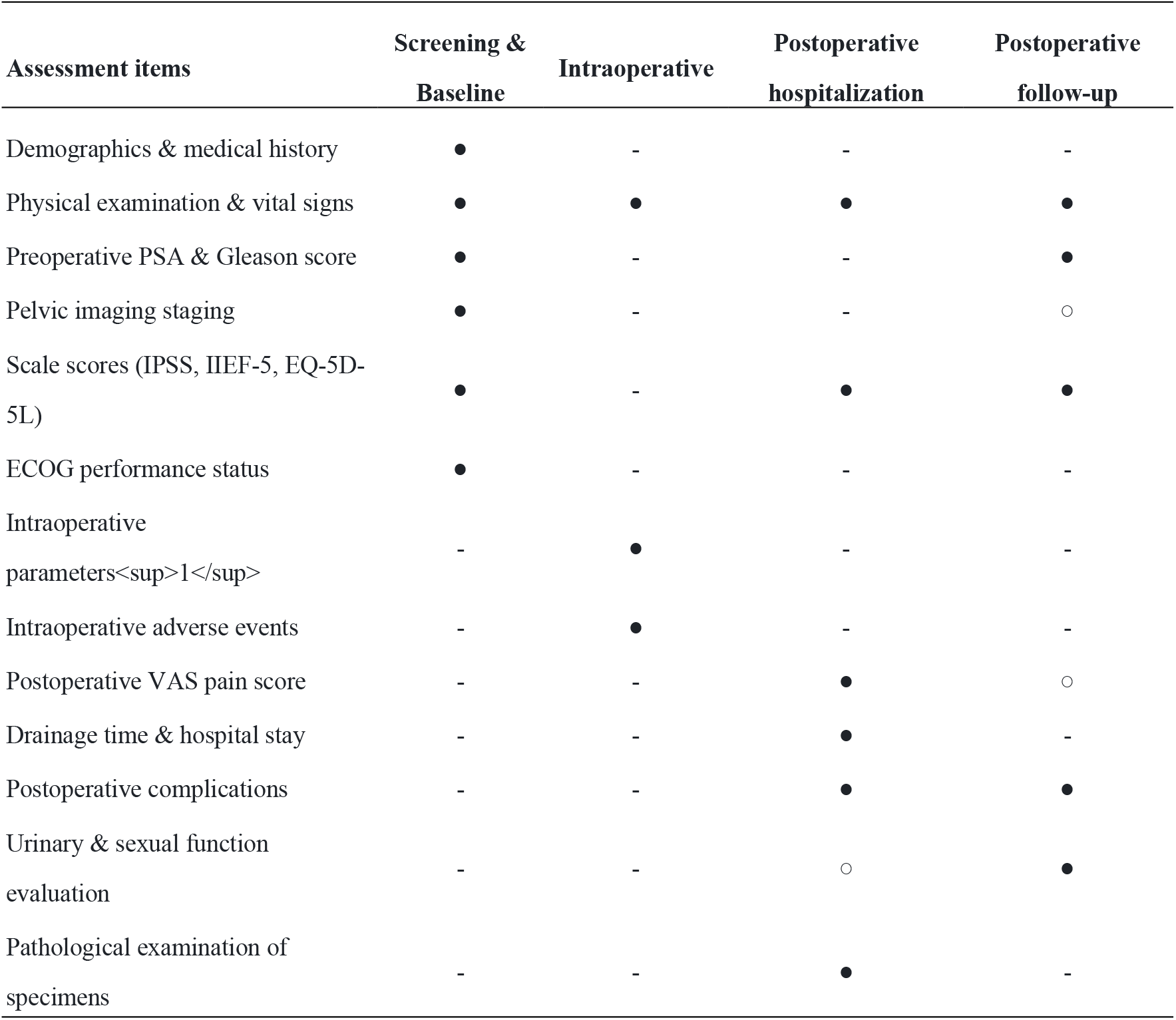
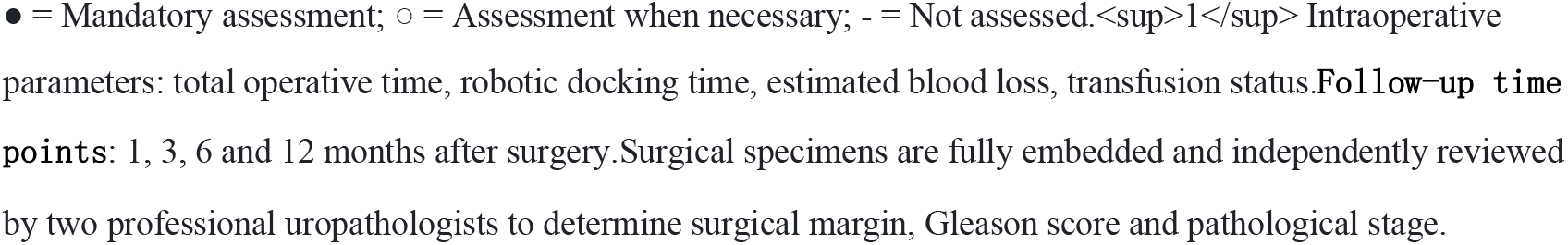
Schedule of data collection and assessments.

| Assessment items | Screening & Baseline | Intraoperative | Postoperative hospitalization | Postoperative follow-up |
| --- | --- | --- | --- | --- |
| Demographics & medical history | ● | - | - | - |
| Physical examination & vital signs | ● | ● | ● | ● |
| Preoperative PSA & Gleason score | ● | - | - | ● |
| Pelvic imaging staging | ● | - | - | ○ |
| Scale scores (IPSS, IIEF-5, EQ-5D-5L) | ● | - | ● | ● |
| ECOG performance status | ● | - | - | - |
| Intraoperative parameters<sup>1</sup> | - | ● | - | - |
| Intraoperative adverse events | - | ● | - | - |
| Postoperative VAS pain score | - | - | ● | ○ |
| Drainage time & hospital stay | - | - | ● | - |
| Postoperative complications | - | - | ● | ● |
| Urinary & sexual function evaluation | - | - | ○ | ● |
| Pathological examination of specimens | - | - | ● | - |

● = Mandatory assessment; ○ = Assessment when necessary; - = Not assessed.<sup>1</sup> Intraoperative parameters: total operative time, robotic docking time, estimated blood loss, transfusion status. **Follow-up time points:** 1, 3, 6 and 12 months after surgery. Surgical specimens are fully embedded and independently reviewed by two professional uropathologists to determine surgical margin, Gleason score and pathological stage.

## 9. Safety monitoring and adverse events

All intraoperative and postoperative adverse events are recorded in detail, especially Clavien-Dindo grade Ⅱ and above events. All serious adverse events (SAEs) will be reported to the hospital ethics committee and clinical trial registry within 24 hours after discovery. The principal investigator and ethics committee will supervise trial safety throughout the study.

## 10. Sample size calculation

This trial is designed as a non-inferiority study based on the primary endpoint (positive surgical margin rate). According to previous literature, the positive margin rate of multiport robotic surgery is estimated at 15%. The non-inferiority margin is set at 7%, one-sided α = 0.025, statistical power (1−β) = 0.80. At least 175 patients are required per group. Considering a 10% dropout rate, the total sample size is 386 (193 cases in each group).

## 11. Statistical analysis

Three analysis sets are defined: intention-to-treat (ITT) set, per-protocol (PP) set and safety analysis set.

Normally distributed continuous variables are presented as mean ± standard deviation and compared using t-test; non-normal data are described as median (interquartile range) and analyzed by rank-sum test. Categorical variables are expressed as number (percentage) and compared with χ^2^ test or Fisher’s exact test. Non-inferiority test is used for the primary outcome. Multiple imputation is applied for missing data. Sensitivity and subgroup analyses are performed as predefined. All analyses are conducted using SPSS 26.0 and R software. A P value < 0.05 is considered statistically significant.

## 12. Ethical considerations

This trial complies with the Declaration of Helsinki and relevant domestic clinical research regulations. All participants sign voluntary informed consent and have the right to withdraw at any time without penalty. All personal health information is protected strictly.

## 13. Data management and quality control

A dedicated research database is established with double data entry and cross-check. All original records are uniformly archived. Regular self-inspection and irregular ethics spot-checks are implemented to ensure standard trial operation.

## 14. Discussion

With the continuous progress of minimally invasive techniques, robotic radical prostatectomy has become the gold standard for the treatment of localized prostate cancer. The extraperitoneal surgical approach is widely adopted in clinical practice, as it avoids peritoneal cavity invasion, lowers the risks of intestinal paralysis, abdominal adhesion and intra-abdominal infection, and well aligns with the enhanced recovery after surgery (ERAS) philosophy. Conventional multiport robotic radical prostatectomy is technically mature, yet multiple abdominal punctures inevitably cause additional soft tissue injury, sustained postoperative pain and unsatisfactory cosmetic effects, which can no longer fully meet patients’ growing demands for ultra-minimally invasive treatment.

Single-port robotic surgery has emerged as a key trend in urological minimally invasive surgery. Using a single operative incision, this technique better preserves abdominal wall integrity, reduces puncture-related trauma and postoperative discomfort, and contributes to faster recovery and improved cosmetic outcomes. Currently, most available data on single-port robotic prostatectomy are based on imported robotic systems. High-quality prospective randomized evidence for domestically developed robotic platforms remains scarce. As a fully independent domestic surgical robot, the KangDuo SR-2000 delivers flexible manipulation, stable intraoperative performance and excellent cost-effectiveness. Its multiport mode has been widely applied in our center with proven reliability, while the oncological efficacy, perioperative safety and functional recovery outcomes of its single-port approach have not been fully validated via controlled trials. This gap has restricted the standardized clinical application of domestic single-port robotic technology.

The present study is designed as a single-center, prospective randomized non-inferiority trial in strict compliance with the SPIRIT 2013 and CONSORT guidelines. We adopted a unified extraperitoneal approach and standardized ERAS protocols across all participants, which effectively eliminates major confounding factors related to surgical access and perioperative management. Postoperative positive surgical margin rate was selected as the primary endpoint to evaluate oncological radical efficacy. A full set of secondary indicators covering perioperative safety, short-term rehabilitation, medium-term urinary and sexual function, and mid-term oncological prognosis was also established, forming a comprehensive evaluation system. If the single-port approach is confirmed to be oncologically non-inferior to the multiport technique, it will offer patients a safer, more comfortable and cosmetically favorable treatment option.

Furthermore, the findings will fill the evidence gap for domestic single-port robotic surgery, provide high-level clinical data to support technical optimization and wider application of the KangDuo system, and facilitate the development of domestic high-end surgical robots.

Several limitations of this trial should be noted. First, this is a single-center study. Despite strict eligibility criteria and standardized procedures, potential selection bias cannot be entirely ruled out. The generalizability of our results needs to be further verified by multi-center, large-sample trials. Second, the scheduled 12-month follow-up period only allows assessment of short-term tumor control and medium-term functional recovery. Long-term oncological outcomes including distant metastasis, cancer-specific survival, as well as late complications and long-term quality of life, require longer follow-up. Third, health economic indicators such as hospitalization costs are not included in this study, and further cost-benefit analysis is warranted. In addition, although a unified ERAS protocol was implemented in this trial, inconsistent execution of ERAS pathways across different medical centers may lead to heterogeneous clinical outcomes in real-world practice.

In conclusion, this rigorously designed randomized trial is of strong clinical practicality. Its results will generate high-level evidence for the application of single-port extraperitoneal robotic radical prostatectomy using the KangDuo system. This study will help optimize individualized minimally invasive strategies for localized prostate cancer, and provide solid clinical evidence for the promotion and technical upgrading of domestic surgical robots. We anticipate that the outcomes will drive the development of urological minimally invasive surgery and the localization of high-end medical devices.

## Data Availability

No datasets were generated or analyzed during the current study. All relevant data from this study will be made available upon study completion. The datasets generated and/or analyzed during the current study will be available from the corresponding author on reasonable request.

